# A highly sensitive XNA-based RT-qPCR assay for the identification of ALK, RET, and ROS1 fusions in lung cancer

**DOI:** 10.1101/2024.01.22.24301177

**Authors:** Bongyong Lee, Andrew Chern, Andrew Y. Fu, Aiguo Zhang, Michael Y. Sha

**Affiliations:** DiaCarta Inc, 4385 Hopyard Rd., Suite 100, Pleasanton, CA 94588

**Keywords:** Lung cancer, Fusion, ALK, RET, ROS1, XNA, RT-qPCR

## Abstract

In lung cancer, the progression of tumors is often fueled by genetic alterations leading to the expression of oncogenic tyrosine kinases. Specifically, chimeric tyrosine kinases involving ALK, RET, and ROS1 are observed in approximately 5-7%, 1-2%, and 1-2% of NSCLS patients, respectively. The presence of these ALK, RET, and ROS1 fusion genes determines the response to tyrosine kinase inhibitors. Consequently, accurate detection of these gene fusions is crucial in the realm of precision medicine. To address this need, we have developed a multiplexed RT-qPCR assay based on xenonucleic acids (XNA) molecular clamping technology for detecting lung cancer fusions. This assay is designed to quantitatively detect thirteen ALK, seven ROS1, and seven RET gene fusions in FFPE samples. The sensitivity of the assay was established at a limit of detection of 50 copies of the synthetic template. Our assay successfully identified all fusion transcripts using 50 ng of RNA from both reference FFPE samples and cell lines. Following this validation, we tested a total of 77 lung cancer patient FFPE samples, demonstrating the effectiveness of our XNA-based fusion gene assay with clinical samples. Notably, this assay is adaptable to highly degraded RNA samples with low input amounts. Our next phase involves expanding the testing to include a broader range of clinical samples as well as cell-free RNAs to further validate its applicability and reliability.

## 1. Introduction

Lung cancer ranks among the most prevalent forms of malignancies and stands as the leading cause of tumor-related fatalities, accounting for nearly a quarter (approximately 25%) of such cases [1]. There are estimated lung cancer new case 117,560 for men and 120,790 for women in 2023 and estimated deaths 67,160 for men and 59,910 for women in 2023 [2]. Over 80% of lung tumors are classified histologically as either adenocarcinomas, squamous cell carcinomas, or large cell carcinomas, collectively categorized as non-small-cell lung cancers (NSCLCs). Various genetic mutations have been identified as drivers of oncogenic processes in NSCLC. These alterations encompass point mutations, deletions, insertions, and gene fusions. In Western nations, approximately 53% of NSCLC cases exhibit mutations in the Kirsten rat sarcoma virus (*KRAS*), epidermal growth factor receptor (*EGFR*), or B-Raf Proto-Oncogene, Serine/Threonine Kinase (*BRAF*) genes. Meanwhile, driver gene fusions and splicing variants are found in 10-15% of patients [3,4]. The prevalent gene fusions observed in NSCLC encompass three genes that encode membrane receptors. These genes are the anaplastic lymphoma receptor tyrosine kinase (*ALK*) with a frequency of 5-7%, the ret proto-oncogene (*RET*) with a frequency of 1-2%, the ROS proto-oncogene 1, receptor tyrosine kinase (*ROS1*) with a similar frequency of 1-2% [5]. The fusion of the echinoderm microtubule-associated protein-like 4 (*EML4*) and the kinesin family member 5B (*KIF5B*) represents significant fusion partners for the *ALK* gene. At least ten different *EML4-ALK* fusion variants and three *KIF5B-ALK* fusion variants have been characterized [6–14]. *ALK* fusions serve as significant predictive biomarkers for tyrosine kinase inhibitors (TKIs). As per the NCCN guidelines, patients with *ALK*-fusion-positive tumors have shown positive responses to ALK TKIs, and alectinib has demonstrated superior efficacy compared to crizotinib when used as the initial treatment option for these patients [15]. *RET* rearrangement was first identified in non-small cell lung cancer (NSCLC) in 2012 using a next generation sequencing assay [16]. Fusions occur when the kinase domain of the *RET* gene combines with the N-terminal region of various gene partners. This fusion leads to the ligand-independent and continuous activation of RET, which in turn stimulates cell proliferation and enhances cell survival [17]. Among these fusion gene partners, the most prevalent partner is *KIF5B*, found in 70-80% of cases, followed by *CCDC6* and *NCOA4* [18]. The FDA has granted approval for two tyrosine kinase inhibitors that specifically target RET, known as selpercatinib and pralsetinib, for the treatment of advanced RET-positive NSCLC [19]. c-ROS oncogene-1 (*ROS1*) gene was first discovered in the 1980s and its role as a proto-oncogene was identified in brain tumors in 2003 [20]. In lung cancer, two *ROS1* fusion transcripts, namely SLC34A2-ROS1 and CD74-ROS1, were initially identified as proto-oncogenes [21]. The rearrangement of *ROS1* primarily occurs in exon 32, 34, 35, or 36, as well as in introns 31 or 33 [22,23]. Among the fusion partners, the most prevalent ones are *CD74* (accounting for 38-54% of cases), followed by *EZR* (13-24%), *SDC4* (9-13%), *SLC34A2* (5-10%), and *GOPC* (2-3%) [23–25]. The *ROS1* rearrangement test is currently advised for all cases of metastatic lung carcinomas. The FDA has granted approval for two tyrosine kinase inhibitors, namely crizotinib and entrectinib, for use as initial treatment options. Although FISH is considered the benchmark method for diagnosing *ALK*, *RET*, and *ROS1* fusions, it is associated with relatively high costs, technical challenges stemming from limited tumor cell availability, and results that can vary based on the operator [26,27]. In contrast, Reverse transcription-quantitative polymerase chain reaction (RT-qPCR), using ROS1 fusion-specific primers, exhibits outstanding performance with a sensitivity of 100% and a specificity of 85.1% [28]. Initially, the identification and characterization of gene fusions in clinical biopsies involved methodologies focused on single alterations. These methods, relying on the polymerase chain reaction (PCR), are still widely used in numerous laboratories. However, the continuously expanding array of targetable fusions prompted the advancement of multiplex techniques. These methods enable the simultaneous investigation of more than one alteration in each assay.

Xenonucleic acid (XNA) is a synthetic DNA analog, originally developed to store genetic information and evolve in response to external stimuli [29]. XNAs have demonstrated remarkable efficiency in binding to specific regular DNA sequences, enabling their utilization as molecular clamps in quantitative real-time PCR or as exceptionally precise molecular probes for identifying specific nucleic acid sequences [30].

The attachment of XNA to its designated sequence obstructs the extension of the DNA strand by DNA polymerase. In cases where there is a mismatch in the target site sequence, the XNA-DNA duplex lacks stability, permitting the extension of the strand by DNA polymerase [31,32].

In this report, we present the development and verification of an innovative multiplex RT-qPCR assay based on XNA. This assay is designed for the simultaneous and qualitative identification of gene fusions that are commonly observed in lung cancer. The fusion biomarker assay covers the detection of fusion events in target genes such as *ALK*, *RET*, and *ROS1*.

## 2. Materials and Methods

### 2.1 Clinical Samples and Cell Lines

Twenty samples preserved in formalin-fixed paraffin-embedded (FFPE) format were supplied by the lung External Quality Assessment (EQA) program conducted by the European Society of Pathology (ESP). Details about the EQA scheme can be found at http://lung.eqascheme.org/info/public/alk/index.xhtml. The organization of ESP schemes adhered to the stipulated requirements for EQA programs in the field of molecular pathology [33]. The clinical information was not provided by the organization. Fifty-seven de-identified FFPE samples from lung cancer patients were obtained from Amsbio (Cambridge, MA, USA). Approval for ethical considerations was granted by the institutional review board at WIRB-Copernicus Group, Inc, and sample collection was conducted with written informed consent. Specimen details are listed in **Supplementary Table S1**. Fusion positive and negative FFPE reference materials were obtained from Horizon Discovery (Cambridge, UK). RNAs were extracted from FFPE sections using the RNeasy FFPE kit (QAIGEN, Hilden, Germany) according to manufacturer’s instructions. Two lung cancer cell lines, A549 (fusion negative cell line) and H2228 (EML4-ALK V3a/b positive cell line) were obtained from American Type Culture Collection (ATCC). CCDC6-RET positive cell line, LC-2/ad was purchased from Millipore Sigma (Burlington, MA, USA). SLC34A2-ROS1 positive cell line, HCC78 was purchased from German Collection of Microorganisms and Cell Cultures (DSMZ). Total RNA was isolated from 1 million cells using a Direct-zol RNA miniprep kit (Zymo Research, Irvine, CA) according to manufacturer’s instructions. Total lung tissue RNA was purchased from Biochain (Newark, CA, USA). RNAs were quantified using the Qubit RNA BR assay kit (Invitrogen, Waltham, MA, USA).

### 2.2 Detection of ALK, RET, or ROS1 Fusion Using RT-qPCR

*ALK*, *RET*, or *ROS1* fusion was detected using the Qfusion^TM^ ALK fusion detection kit, Qfusion^TM^ RET fusion detection kit, or Qfusion^TM^ ROS1 fusion detection kit (DiaCarta Inc., Pleasanton, CA, USA), which can simultaneously detect thirteen *ALK* fusions, seven *RET* fusions, or seven *ROS1* fusions, respectively (**Supplementary Figure S1**). Briefly, 50 ng of RNAs were subjected to RT-qPCR using Bio-Rad CFX384 (Bio-Rad Laboratories, Inc., Hercules, CA, USA). The thermal cycling condition was 10 min at 50 °C for reverse transcription (RT), 2 min at 95 °C for RT inactivation, followed by 45 cycles of 5 sec at 95 °C for denaturation, 40 sec at 70 °C for XNA binding, 30 sec at 64 °C for primer binding and elongation with slow ramp rate (3 °C per second).

### 2.3 Determination of Analytical Sensitivity

The analytical sensitivity was assessed using synthetic double-stranded DNAs (**Supplementary Table S2**). gBlocks gene fragments were purchased from Integrated DNA Technologies (Coralville, IA, USA). Three distinct copy numbers (500, 100, and 50 copies) of gBlocks were employed as inputs, and the RT-qPCR assay was conducted with 18 replicates.

For assessing the limit of detection of RNA fusion targets in cell lines, a total RNA input of 50 ng was employed. The total RNA from each cell line underwent serial dilution with normal lung tissue RNA (Biochain) and was subsequently assayed using the Qfusion^TM^ ALK fusion detection kit, Qfusion^TM^ RET fusion detection kit, or Qfusion^TM^ ROS1 fusion detection kit (DiaCarta Inc.). Similarly, for RNA fusion targets in the FFPE RNA reference standard (Horizon Discovery), RNAs were diluted with normal lung tissue FFPE RNA (Horizon Discovery) and assayed using the Qfusion^TM^ assays.

### 2.4 Detection of ALK, RET, or ROS1 Fusion Using Next Generation Sequencing (NGS)

Lung cancer specific fusions were analyzed using a OptiSeq^TM^ lung cancer fusion NGS panel (DiaCarta Inc., Pleasanton, CA, USA). Fifty nanograms of RNAs underwent library preparation following the manufacturer’s guidelines. The libraries were subsequently sequenced on an Illumina MiSeq platform (Illumina, San Diego, CA, USA) with read lengths of 150 bases, generating approximately 0.1 million paired-end reads per library. The obtained results were analyzed using the QIAGEN CLC Genomics Workbench version 20.0.4 (QIAGEN, Aarhus, Denmark) for fusion detection. A fusion event was considered confirmed if a minimum of 10 reads were identified as crossing the fusion junction.

### 2.5 Preparation of Fusion and Wild type Transcripts by In Vitro Transcription

EML4-ALK V1 and wild-type ALK transcripts were synthesized using the Megascript T7 transcription kit (Thermo Fisher Scientific, Carlsbad, CA, USA) following the manufacturer’s guidelines. Subsequently, RNAs were purified using the RNA Clean & Concentrator-5 kit (Zymo Research) and quantified with the Qubit RNA BR assay kit (Invitrogen).

## 3. Results

### 3.1. Analytical Sensitivity, Limit of Detection (LoD)

To assess the analytical sensitivity of each Qfusion^TM^ assay designed for detecting *ALK*, *RET*, or *ROS1* fusions, we conducted 18 replicate reactions with 500, 100, and 50 copies of synthetic double-stranded fusion templates (as outlined in **Supplementary Table S2**). The analytical sensitivity was expressed as the number of copies detected per reaction. The results demonstrated a 100% detection rate when testing with 500, 100, and 50 copies per reaction for most templates. However, there was a slightly lower detection rate of 89% for the EML4-ALK V6 template and 83% for the GOPC-ROS1 G8;R25 template (**Table 1a-c**). Therefore, the limit of detection for all three detection assays was established at 50 copies of the target per reaction.

**Table 1.**
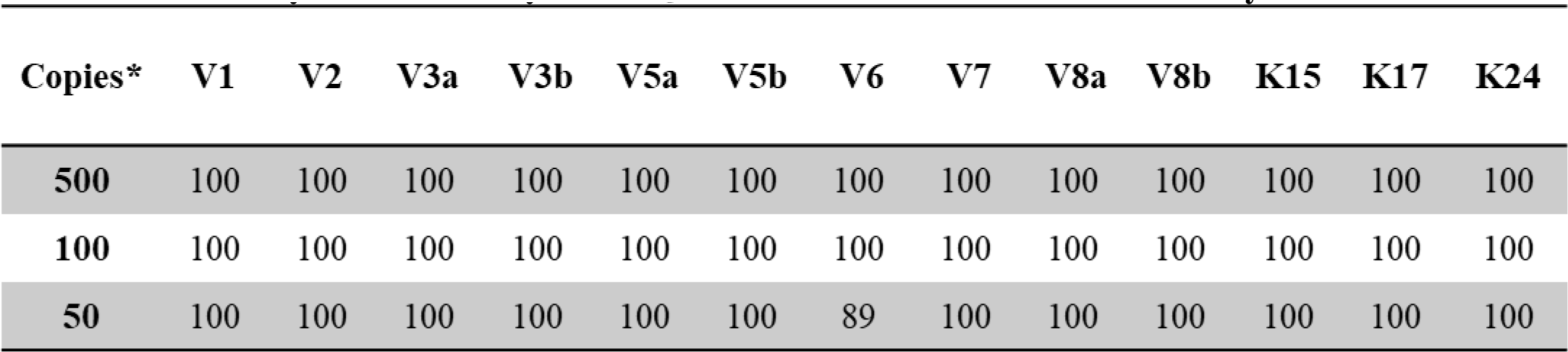
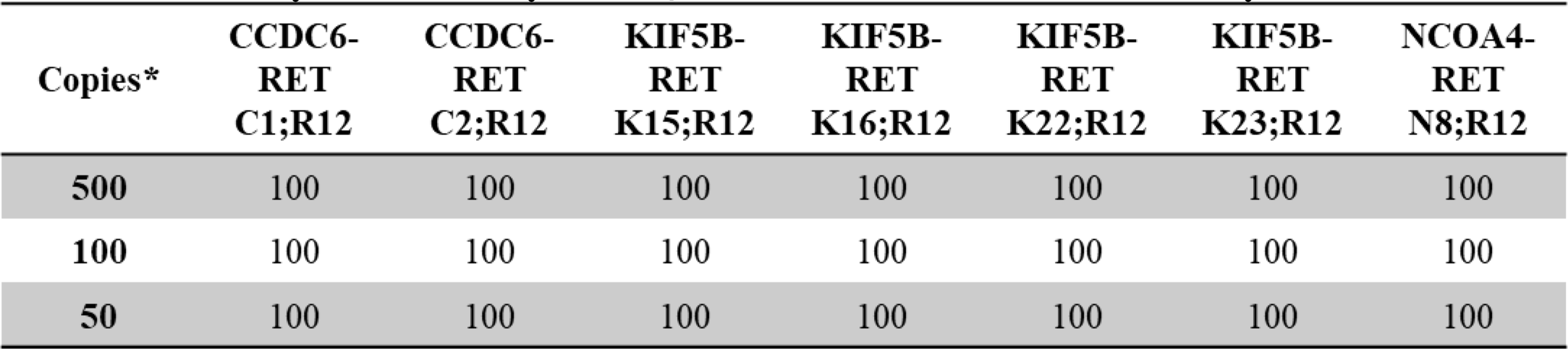
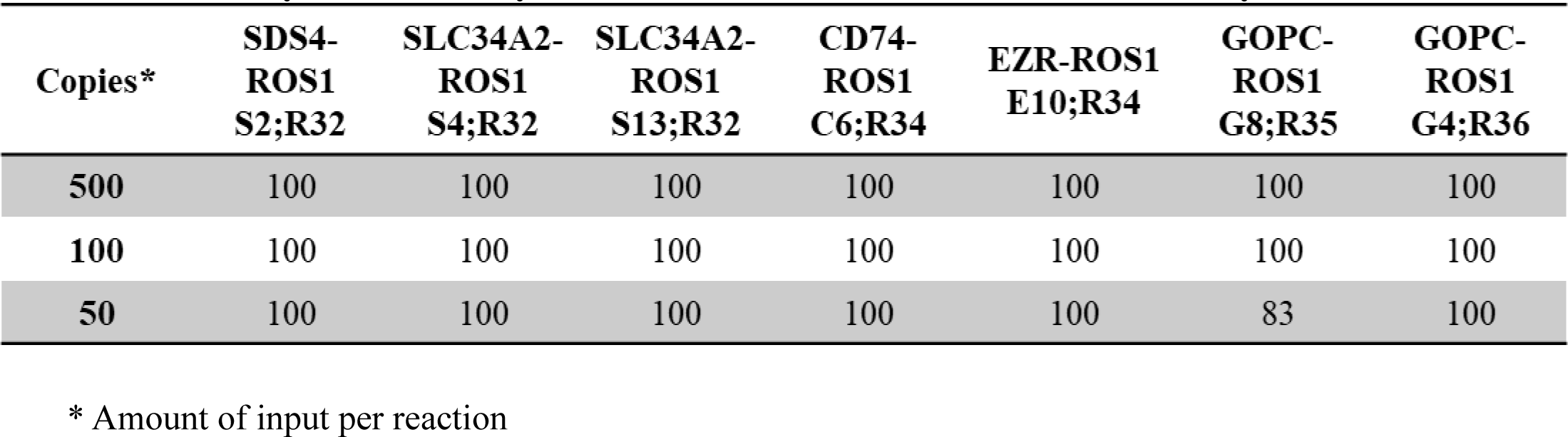
a. Analytical Sensitivity of the Qfusion^TM^ ALK Fusion Detection Assay b. Analytical Sensitivity of the Qfusion^TM^ RET Fusion Detection Assay c. Analytical Sensitivity of the Qfusion^TM^ ROS1 Fusion Detection Assay

### 3.2 Enhancement of Qfusion^TM^ Assay Specificity and Sensitivity with XNA

In multiplex PCR-based assays, off-target amplification is a common issue [34]. Particularly, in the presence of a substantial background of wild-type sequences, false positives in mutation or fusion detection tend to increase, primarily due to the promiscuous binding of primers and probes to sequences that closely resemble the target [31]. To mitigate non-specific amplification in the presence of abundant wild-type backgrounds, we assessed the detection efficiency of each Qfusion^TM^ assay by employing 10-fold serial dilutions of ALK, RET, or ROS1 wild-type RNA transcripts. As illustrated in **Figure 1a**, each assay exhibited non-specific detection of wild-type RNA transcripts. However, the presence of XNA efficiently suppressed the detection of wild-type RNAs. This suggests that XNA enhances the assay’s specificity, thereby reducing the occurrence of false positives.

**Figure 1.**
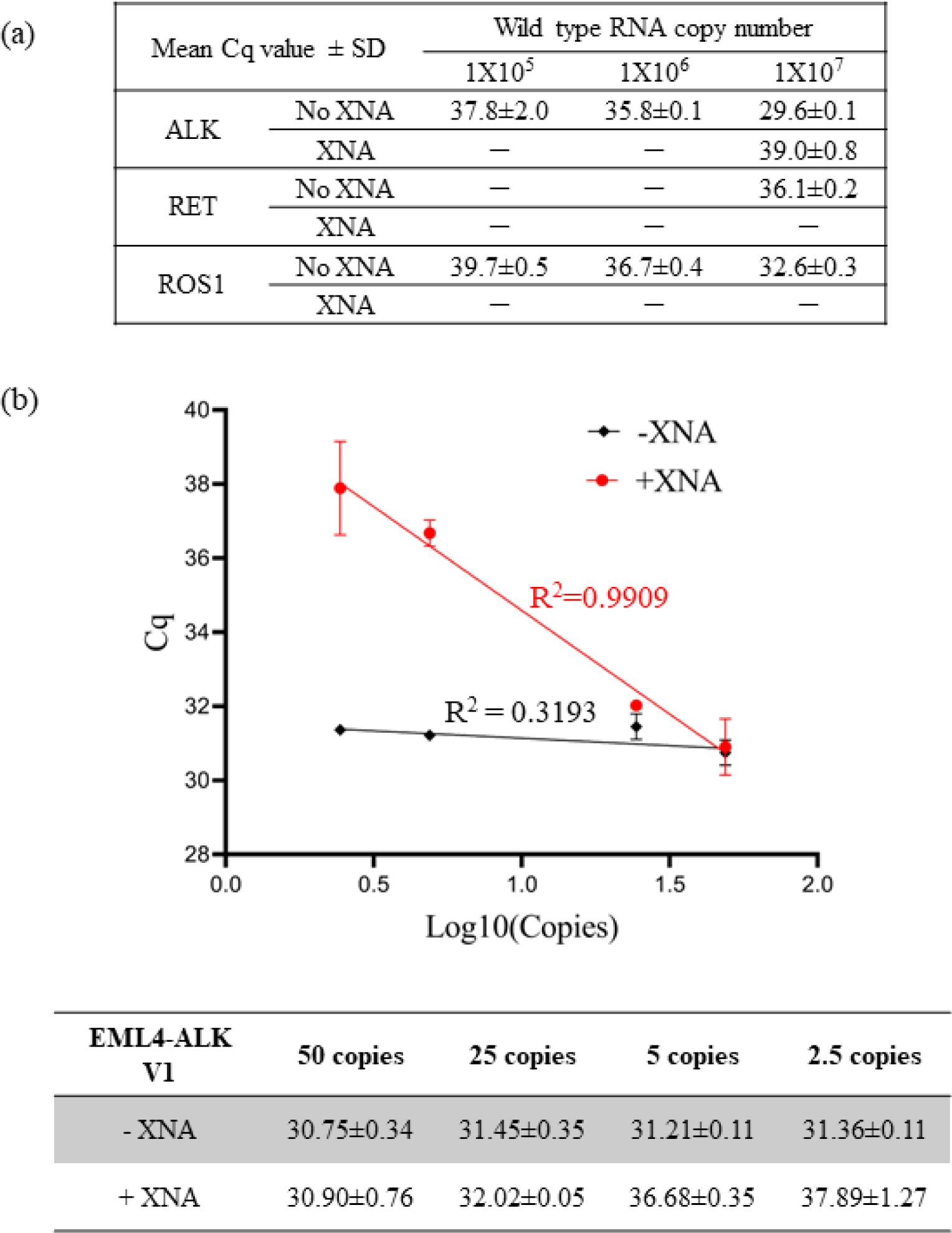
Enhancing Assay Specificity and Sensitivity with XNA. (a) XNA enhanced specificity by preventing non-specific amplification of abundant wild-type sequences. (b) XNA improved assay sensitivity by inhibiting the wild-type backgrounds.

To assess the influence of XNA on assay sensitivity, we conducted multiple measurements using varying quantities of EML4-ALK V1 synthetic DNA fragments, both in the presence and absence of 10,000,000 copies of wild-type ALK DNA fragments. Given that the presence of abundant wild-types could lead to non-specific amplification in the reaction (**Figure 1a**), we hypothesized that if XNA impeded the amplification of abundant wild-type ALK, it would enhance the detection limit of the fusion target by eliminating non-specific amplification of wild-types. Furthermore, we postulated that XNA would not impact the detection of fusion transcripts in the absence of wild-type ALK.

As anticipated, our results affirmed that XNA had no significant impact on the detection of EML4-ALK V1 fusion (**Supplementary Table S3**). As depicted in **Figure 1b**, in the absence of XNA, the Cq value remained unchanged for different copy numbers of fusion targets due to preferential amplification of abundant wild types, masking the amplification of fusion targets. Consequently, at a given level of fusion inputs, the assay was unable to distinguish between 50 copies and 2.5 copies, resulting in a 50-copy LoD. However, XNA effectively prevented the amplification of wild-types, enabling the detection of low copy numbers of fusion targets in a linear, dose-dependent manner ranging from 2.5 to 50 copies. This implies that the LoD was enhanced to the 2.5-copy level, demonstrating increased sensitivity.

In summary, XNA inhibited the amplification of wild-types, thereby enhancing the specificity and sensitivity of the Qfusion^TM^ assay.

### 3.3 Evaluation of the Qfusion^TM^ Assays Performance with Cell Lines and FFPE RNA Reference Standards

The sensitivity and specificity of individual Qfusion^TM^ assays were subsequently assessed through examination of fusion-negative and positive cancer cell lines. RNA extraction was performed on four cell lines: A549 (fusion-negative), H2228 (EML4-ALK V3/b positive), LC-2/ad (CCDC6-RET C1;R12 positive), and HCC78 (SLC34A2-ROS1 S4;R32 positive). Dilution of RNAs from cancer cell lines was carried out with normal lung tissue RNA, ranging from 100% to 0% representation of cancer portions. Through the systematic dilution of cancer cell line RNAs, the sensitivity and specificity of the Qfusion^TM^ assay were assessed. The individual XNA-mediated ALK, RET, or ROS1 Qfusion^TM^ assays successfully identified all correct fusion targets at a minimum dilution of 1%, with corresponding Cq values of 33.62±0.50, 27.03±0.13, and 25.42±0.10, respectively. Notably, no fusion signals were detected in the A549 fusion-negative cell line (**Figure 2a**).

**Figure 2.**
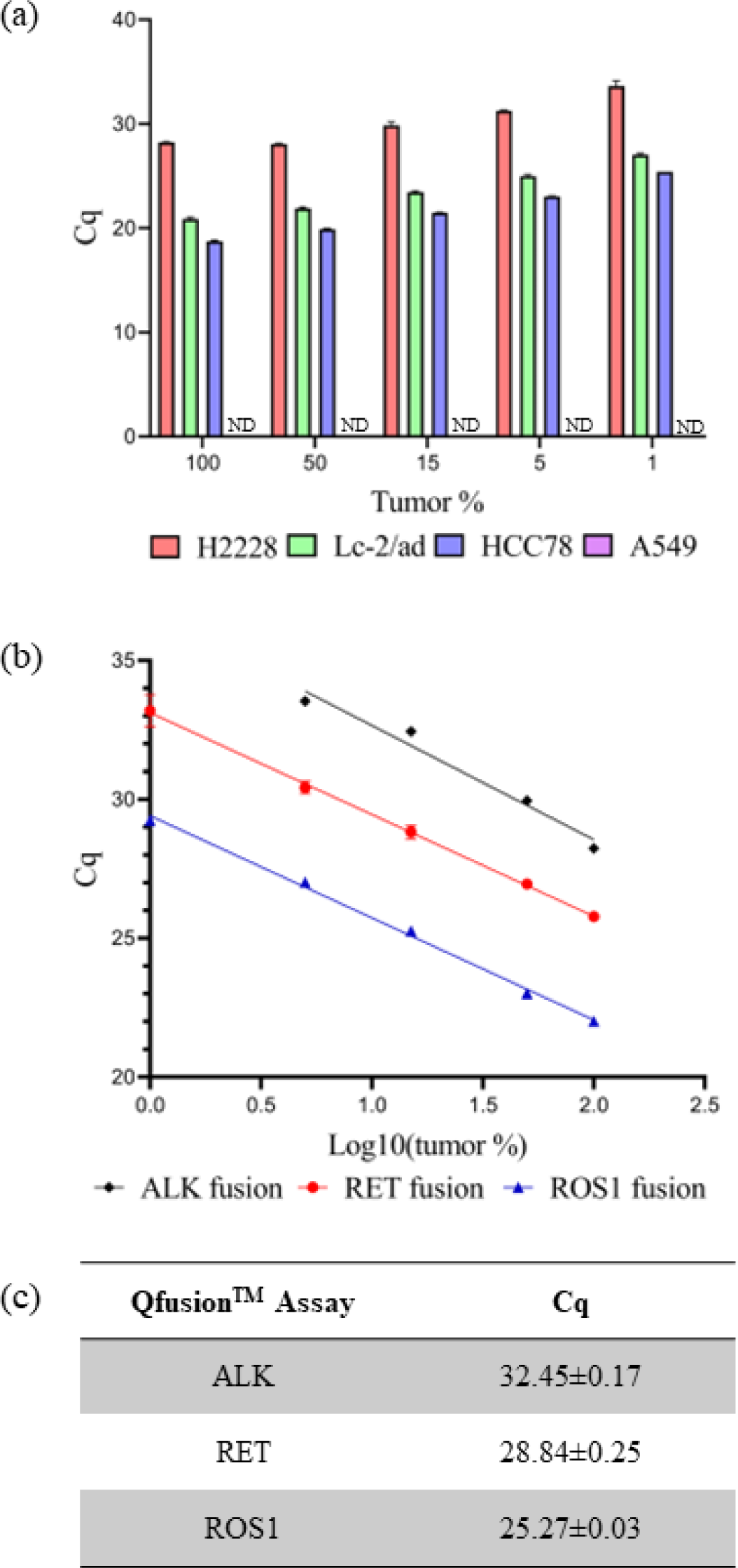
Evaluating Qfusion^TM^ assay performance. (a) Each Qfusion^TM^ assay was tested with corresponding cell line: A549, H2228 (EML4-ALK), LC-2/ad (CCDC6-RET), and HCC78 (SLC45A-ROS1). A549 was fusion negative cell line and Cq was not determined (ND). (b, c) FFPE RNA reference standard was diluted with normal FFPE RNA and analyzed with each Qfusion^TM^ assay. Cq values for 15% tumor samples were determined.

The current gold standard diagnostic method for detecting *ALK*, *RET*, or *ROS1* fusion is Fluorescence *In Situ* Hybridization (FISH). A positive fusion result is typically determined if at least 15% of tumor cells exhibit rearrangement [35]. Given that the Qfusion^TM^ assay was specifically designed to detect fusions in both archived and fresh tissue samples, the Cq values for each Qfusion^TM^ assay at the 15% tumor cell threshold were established. To ascertain these Cq values, RNAs were prepared from commercially available Formalin-Fixed Paraffin-Embedded (FFPE) reference standards and normal lung FFPE samples. Various concentrations of reference standard FFPE RNAs, when diluted with normal lung FFPE RNAs, underwent analysis using the Qfusion^TM^ assay. The resulting Cq values for the Qfusion^TM^ ALK, RET, or ROS1 fusion assays at a 15% tumor fraction were determined as 32.45±0.17, 28.84±0.25, and 25.27±0.03, respectively (**Figure 2b & 2c**).

### 3.4 Evaluation of the Qfusion^TM^ Assays Performance Using Clinical Samples

In our previous study, we examined twenty FFPE patient samples acquired from the EQA program of the ESP using the OptiSeq^TM^ lung cancer fusion NGS panel [36]. This panel, specifically designed for the detection of 63 known lung cancer-specific fusion genes, including *ALK*, *RET*, and *ROS1* fusion genes, successfully identified SLC45A-ROS1 fusions in two patient samples (**Figure 3a**), with no instances of ALK or RET fusions. Subsequently, the same twenty samples were subjected to testing with the Qfusion^TM^ ALK, RET, or ROS1 fusion detection assay. As illustrated in **Figure 3b**, the Qfusion^TM^ ROS1 fusion detection assay identified the same patients as ROS1 fusion-positive. Conversely, both the Qfusion^TM^ ALK and RET fusion detection assays indicated that all tested samples were negative for *ALK* or *RET* fusion.

**Figure 3.**
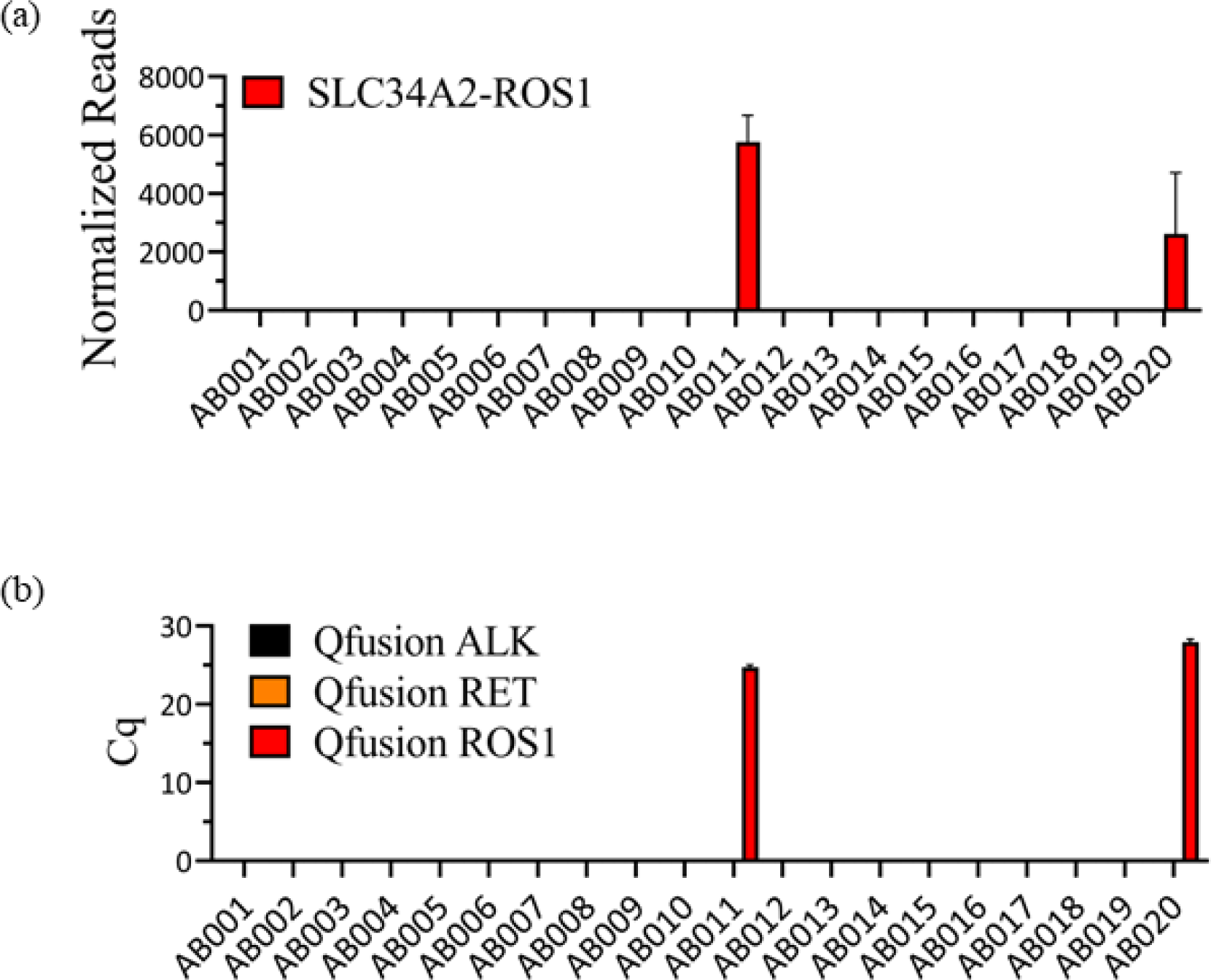
Consistency Between OptiSeq^TM^ Lung Cancer Fusion NGS Panel and Qfusion^TM^ ALK, RET, or ROS1 Fusion Detection Assay Results. (a) Analysis of twenty FFPE samples using the OptiSeq^TM^ lung cancer fusion NGS panel revealed the presence of SLC34A2-ROS1 fusion in two patient samples. The Y-axis represents normalized reads, and error bars indicate the standard deviation of three replicates. (b) The same twenty FFPE samples were subjected to analysis using Qfusion^TM^ assays. The results confirmed ROS1 fusion positivity in two patients with SLC34A2-ROS1 fusion. Error bars depict the standard deviation of three replicates.

The consistent findings observed between the Qfusion^TM^ assays and the NGS assay prompted us to extend our investigation to a larger set of patient samples. The Qfusion^TM^ ALK, RET and ROS1 fusion detection assay was employed to assess 57 distinct clinical FFPE samples obtained from Amsbio (Cambridge, MA, USA). Within the 57 samples, one patient was identified as *ALK* fusion-positive, one as *RET* fusion-positive, and one as *ROS1* fusion-positive (**Figure 4a**). Notably, one patient sample, AMS029, exhibited concurrent *ALK* and *RET* fusions. To validate the outcomes derived from the Qfusion^TM^ assay and to determine the specific fusion genes implicated, all samples identified as fusion-positive underwent a reanalysis employing individual PCR amplicon. The Qfusion^TM^ assay product (amplicon) served as the template for the second individual PCR, wherein a single primer pair and probe were employed to discern the specific fusion variant.

**Figure 4.**
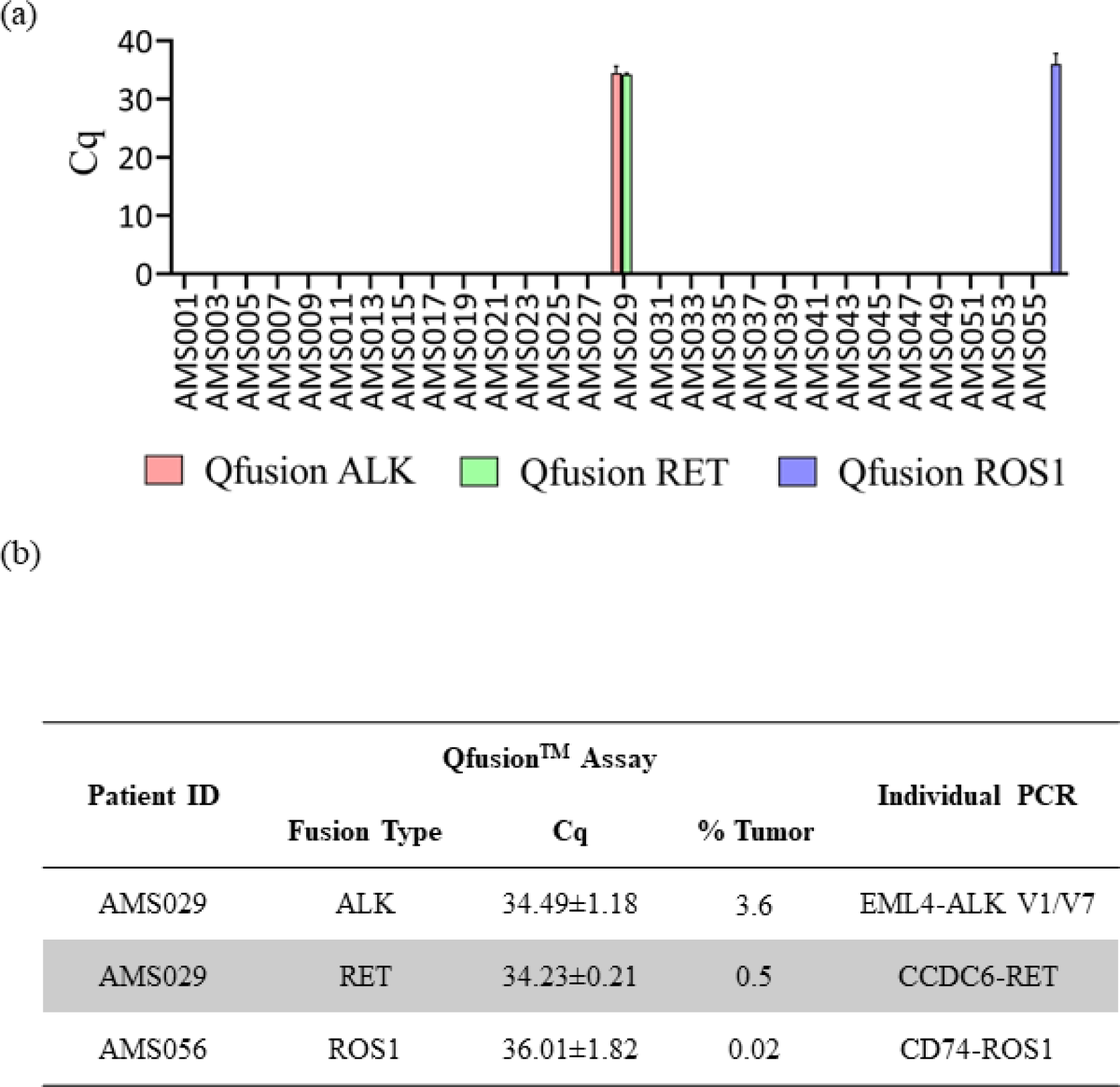
Assessment of Qfusion^TM^ assays performance utilizing clinical samples. (a) Results from the analysis of 57 FFPE lung cancer patient samples using the Qfusion^TM^ ALK, RET, or ROS1 fusion detection assay. The data reveals the identification of one patient with ALK and RET fusions, and one patient with ROS1 fusion. (b) Validation of distinct fusion events through singleplex RT-qPCR. The identified ALK fusions included EML4-ALK V1/V7, while the RET fusion involving CCDC6-RET. Additionally, a single ROS1 fusion event, CD74-ROS1, was confirmed.

The singleplex individual PCR revealed that the AMS029 patient harbored two types of EML-ALK fusions, V1 and V7 variants, along with one CCDC6-RET fusion. In the case of the ROS1 fusion-positive AMS056 patient, the singleplex individual PCR identified the CD74-ROS1 fusion variant (**Figure 4b**). Subsequently, each individual PCR product underwent analysis through Sanger sequencing, thereby confirming the presence of the corresponding fusion variants (**Supplementary Figure S2**).

## 4. Discussion

Chromosomal translocations resulting in the formation of fusion genes represent a significant causative factor in cancer, and the precise diagnosis of these events is crucial for effective treatment. The prevailing diagnostic method, FISH, relies on pre-existing annotations, exhibiting low throughput and limited resolution. In contrast to FISH, RNA NGS sequencing (RNAseq) provides a high-resolution approach to detecting fusion genes, capable of identifying both established and novel fusions. While the high-throughput nature of RNAseq expands diagnostic possibilities, it also introduces a challenge in terms of increased false-positive rates, indicating potential inaccuracies in fusion gene calls and the identification of non-driver fusion events, such as recurrent chimeric fusion RNAs in non-cancerous tissues and cells [37].

Moreover, incorporating RNAseq into routine clinical practice for primary fusion detection is impractical due to its elevated cost, prolonged turnaround time, and intricate procedural and data analysis requirements. Therefore, our focus has shifted towards the development of a cost-effective, high-throughput fusion detection assay utilizing a multiplex-based reverse RT-qPCR approach.

We harnessed our existing XNA technology, previously employed for Single Nucleotide Polymorphism (SNP) detection in genomic DNA samples [31], to design assays for the identification of gene fusion transcripts. Specifically, we formulated panels targeting 28 well-known fusion transcripts, encompassing over 90% of ALK, RET, or ROS1 fusion transcripts [38]. The assays were systematically tested on *in vitro* transcribed RNA, cell lines, and FFPE samples. In each instance, we successfully identified both known fusion transcripts and fusions in patient FFPE samples with previously unknown fusion status.

The inherent characteristics of multiplex PCR, including diminished specificity and sensitivity compared to singleplex PCR, are evident [39,40]. As illustrated in **Figure 1**, non-specific amplification of wild-type ALK transcripts was observed despite employing fusion-specific primer and probe sets. However, the introduction of XNA efficiently mitigated this non-specific amplification increasing assay specificity and sensitivity. Our utilization of XNA technology, originally employed for SNP detection on DNA, has been extended to include fusion detection on RNA.

All three Qfusion^TM^ assays demonstrated the detection of synthetic targets at levels as low as 50 copies per reaction, with 18 replicate measurements. When evaluating analytical sensitivity using cell lines and FFPE RNA references, the assays reliably identified corresponding targets at tumor fractions as low as 1%. However, it is noteworthy that the fusion positivity criteria were established based on FISH standards, where a 15% tumor cell population with fusion is considered a positive case.

When applying this criterion to 57 fusion-unknown patient FFPE samples, we identified two fusion-positive cases, each exhibiting a frequency of 1.8% for ALK fusion, RET fusion, and ROS1 fusion, respectively. Interestingly, one patient displayed two distinct fusions, ALK and RET fusions, a finding confirmed by singleplex PCR. The occurrence of such dual fusions in a single patient is considered rare and may be attributed to the inherent nature of intra-tumor heterogeneity [41]. The fusion frequency we observed in this study, mirroring the general low frequency of fusion-positive cases among lung cancer patients, reinforces the challenges posed by the relative rarity of these events in the broader patient population. This insight contributes to our understanding of the prevalence of fusion-positive cases in lung cancer and underscores the importance of sensitive and comprehensive diagnostic approaches for detecting such infrequent but clinically significant occurrences.

Enabling the timely and precise identification of actionable biomarkers is a crucial advancement in making testing more widely accessible. In this regard, we present RT-qPCR panels that comprehensively address actionable targets, showcasing a notable level of agreement with NGS results. The streamlined workflow and straightforward analysis associated with these panels offer a promising solution to improve the accessibility of biomarker testing, thereby contributing to the advancement of personalized medicine guidance.

## Data Availability

All data produced in the present work are contained in the manuscript

