## Supplementary table and figure for "A highly sensitive XNA-based RT-qPCR assay for the identification of ALK, RET, and ROS1 fusions in lung cancer"

**Supplementary Table and Figure Legends**

**Table S1. Amsbio FFPE sample information**

**Table S2. Sequence information of synthetic fusion and wild type templates**

**Table S3. The effect of XNA on fusion detection**

**Figure S1. Schematic of Qfusion^TM^ ALK, RET, or ROS1 fusion detection assay targets.**

**Figure S2. Confirmation of fusion genes by Sanger sequencing.**

Red arrow indicates the breaking point.

**Table S1**. Amsbio FFPE sample information
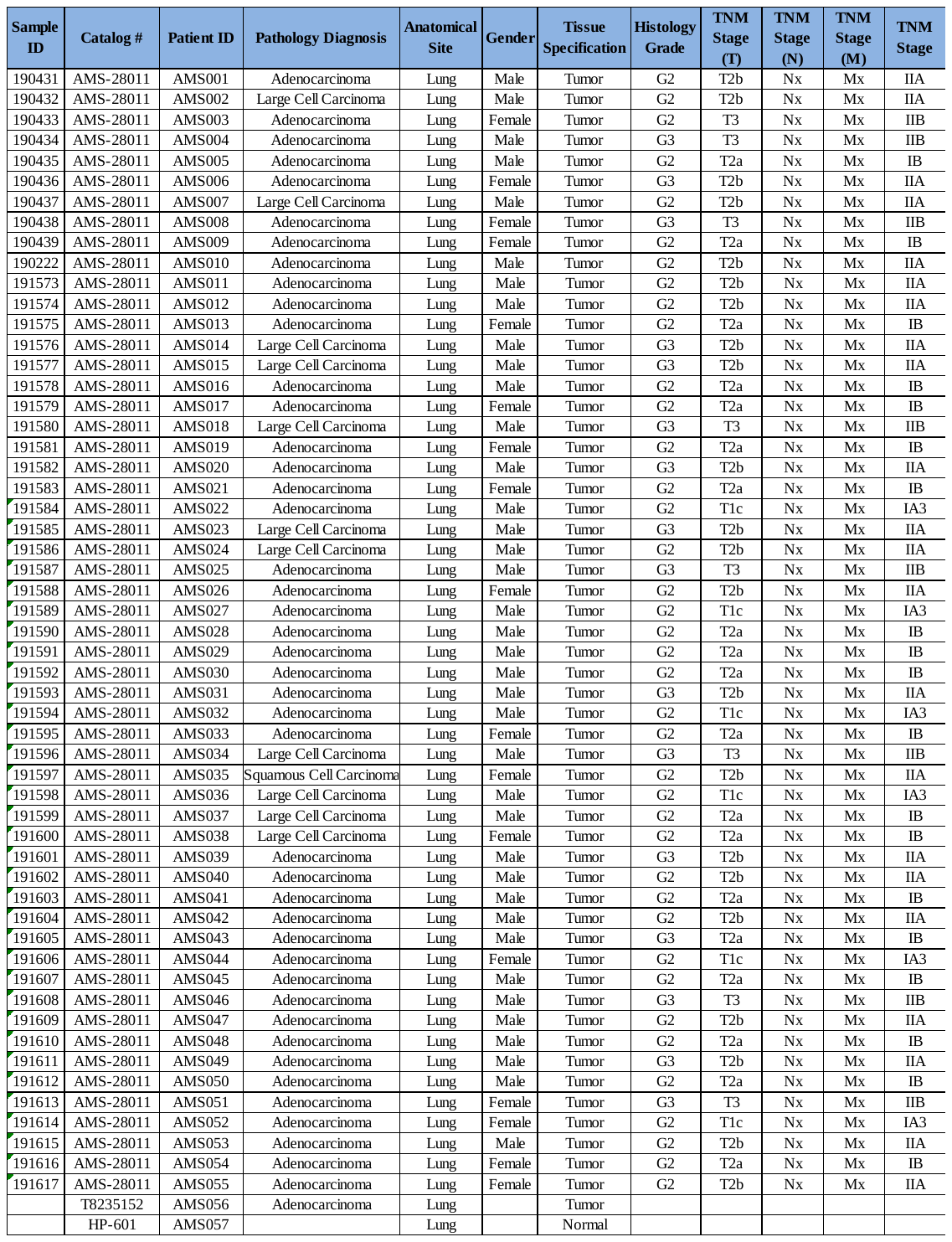

**Table S2**. Sequence information of synthetic fusion and wild type templates

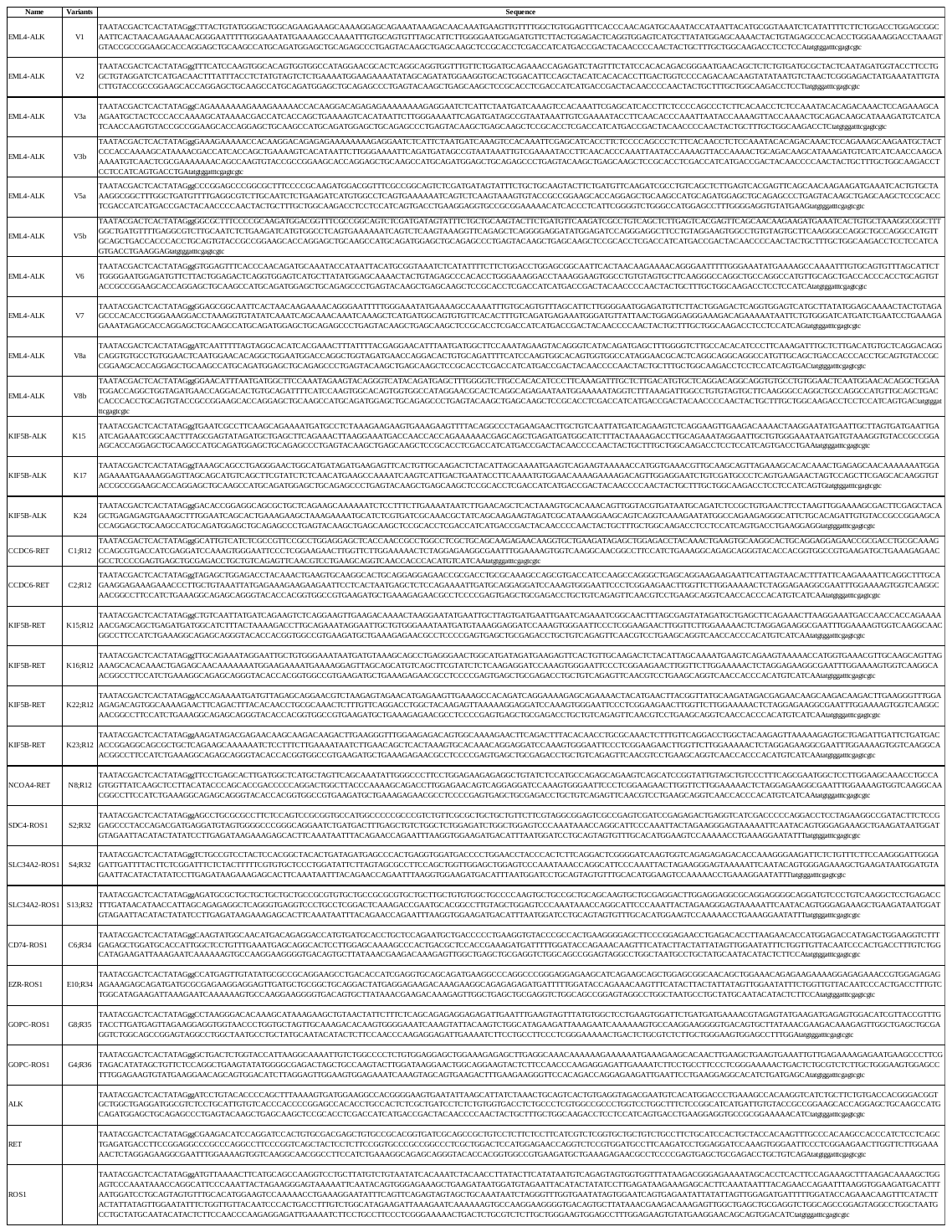

**Table S3**. The effect of XNA on fusion detection

| **EML4-ALK V1** | **50 copies** | |
| --- | --- | --- |
|  | no XNA | XNA |
| **Cq Rep 1** | 31.04 | 31.57 |
| **Cq Rep 2** | 30.69 | 30.84 |
| **Cq Rep 3** | 30.86 | 31.14 |
| **Cq Rep 4** | 31.01 | 30.95 |
| **Cq Rep 5** | 31.04 | 31.12 |
| **Cq Rep 6** | 31.10 | 31.15 |
| **Cq Rep 7** | 30.99 | 31.36 |
| **Cq Rep 8** | 30.99 | 31.25 |
| **Cq Rep 9** | 31.06 | 31.35 |
| **Cq Rep 10** | 30.84 | 31.15 |
| **AVG** | 30.96 | 31.19 |
| **SD** | 0.13 | 0.21 |
| **Delta Cq** |  | 0.22 |

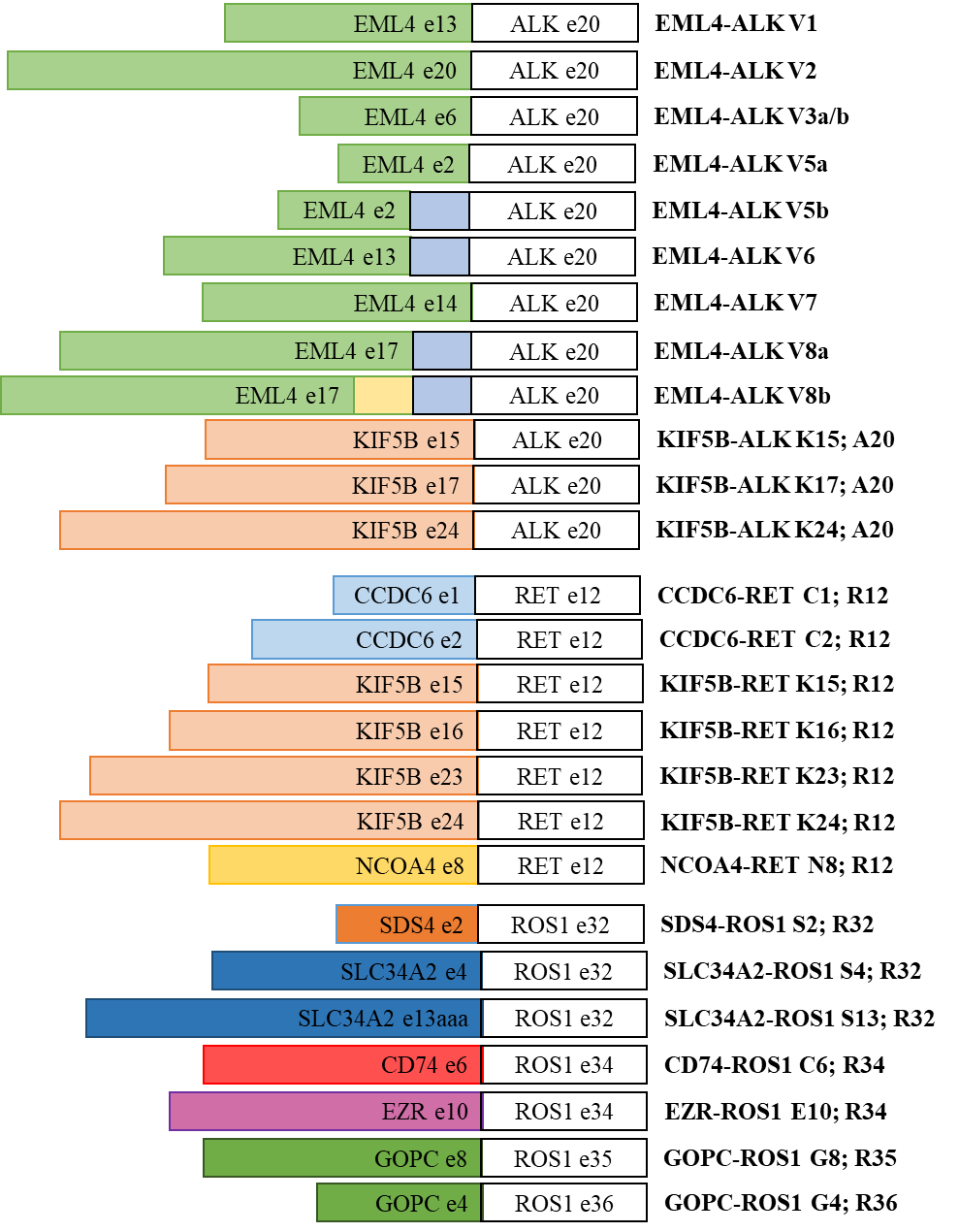
**Figure S1. Schematic of Qfusion^TM^ ALK, RET, or ROS1 fusion detection assay targets.**

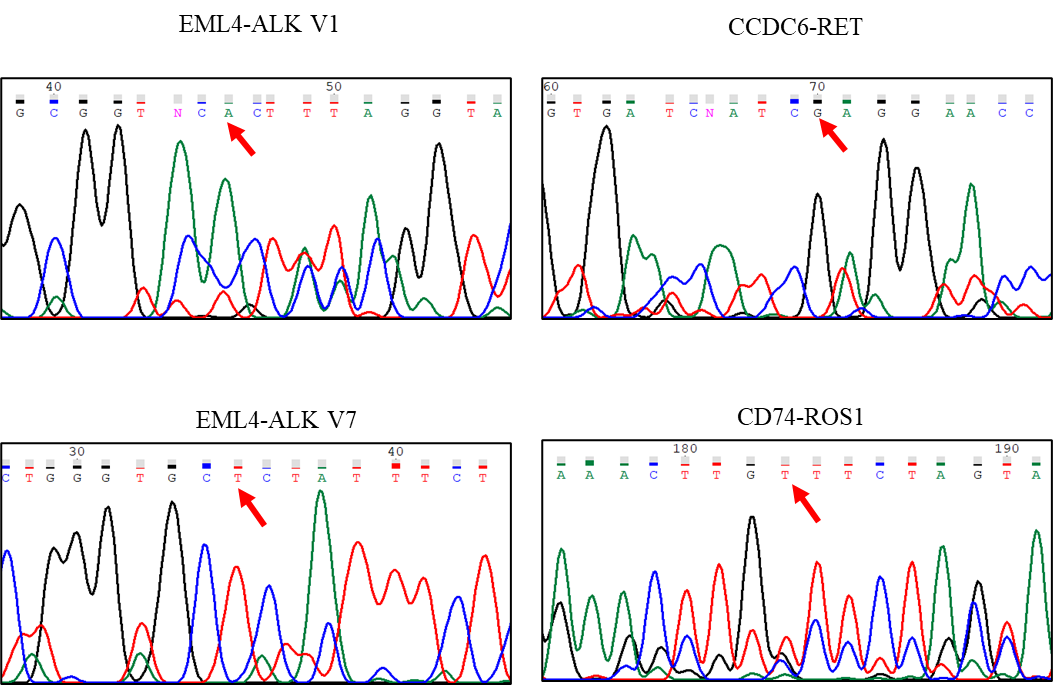

**Figure S2. Confirmation of fusion genes by Sanger sequencing.**

Red arrow indicates the breaking point.
